# Success in vaccination programming through community health workers: A case study of Nepal, Senegal, and Zambia

**DOI:** 10.1101/2023.05.05.23289567

**Authors:** Emily Ogutu, Anna S. Ellis, Kyra A. Hester, Katie Rodriguez, Zoe Sakas, Chandni Jaishwal, Chenmua Yang, Sameer Dixit, Anindya S. Bose, Moussa Sarr, William Kilembe, Robert A. Bednarczyk, Matthew C. Freeman

## Abstract

**Introduction:** Community health workers are essential to frontline health outreach throughout low- and middle-income countries, including programming for early childhood immunization. The World Health Organization estimates a projected shortage of 18 million health workers by 2030. Understanding how community health workers are engaged for successful early childhood vaccination among countries who showed success in immunization coverage would support evidence-based policy guidance across contexts. To that end, we identified factors of community health worker programs that contributed to improved vaccination coverage in Nepal, Senegal, and Zambia.

**Methods:** We conducted interviews and focus group discussions at the national, regional, district, health facility, and community levels of the health systems of Nepal, Senegal, and Zambia, and used thematic analysis to investigate contributing factors of community health worker programming that supported early childhood immunization within each country and across contexts. We developed a model that could be used for assessment and adaptation based on lessons learned.

**Findings:** Across all countries, implementation of vaccination programming relied principally on the 1) organization, 2) motivation, and 3) trust of community health workers. Organization was accomplished by expanding cadres of community health workers to carry out their roles and responsibilities related to vaccination. Motivation of community health workers was supported by intrinsic and extrinsic incentives. Trust was expressed by communities due to community health worker respect and value placed on work.

**Conclusion:** Improvements in immunization coverage followed successful community health worker programs, facilitated by diversification of cadres, roles and responsibilities, motivation, and trust. With the continued projection of health worker shortages, especially in low-income countries, community health workers bridged the equity gap in access to vaccination services by enabling wider reach to minority populations and populations in hard-to-reach areas. Although improvements in vaccination programming were seen in all three countries - including government - commitment to addressing human resource deficits, training and renumeration; workload, low and inconsistent compensation, inconsistency in training duration and scope, and supervision are still major challenges to immunization programming. Vaccination and health decision-makers should consider organization, motivation, and trust of community health workers to improve the implementation of immunization programming.

## Introduction

Despite efforts to increase early childhood immunization over the past 20 years, many countries continue to report excessive morbidity and mortality from vaccine-preventable diseases. Maintaining high vaccination coverage is of economic benefit by preventing early childhood disease, improving health outcomes, and reducing expenditure on treatment[1, 2]. The Immunization Agenda 2030 (IA2030) aims to maintain hard-won gains in immunization by leaving no one behind at any stage of life through increasing equitable access and use of new and existing vaccines, and strengthening immunization within primary healthcare. Through this strategy, countries define their own targets and timelines to achieve IA2030 goals[3]. Achieving this goal requires strategizing to address multiple inefficiencies, including health system strengthening, infrastructural development and increased human resources.

The 2030 targets for the Global Strategy for Human Resources for Health, developed for low and middle-income countries, are to create, fill, and sustain at least 10 million additional jobs in the health and social care sectors to address unmet needs for the equitable and effective coverage of health services[4]. The World Health Organization (WHO) estimates a projected shortage of 18 million health workers by 2030, mostly in low- and lower-middle-income countries[5]. Estimates include the need for 44.5 basic health workers (e.g., doctors, nurses, and midwives) per 10,000 population; however, only half of the WHO member states have that level at present, with Southeast Asia and sub-Saharan Africa having the greatest absolute shortage of health workers[4]. Community health workers (CHWs)- are community members who volunteer to serve as frontline healthcare workers in underserved communities to bridge the human resource deficit[6]. Continued expansion of the numbers, functions, training, and support of CHWs and provision of financial packages commensurate to their job demands and workload could bridge the gap because of the feasibility of CHWs deployment and effectiveness[7, 8].

As frontline health workers, CHWs play a critical role in routine vaccination programming by generating demand for childhood vaccination; connecting community members to the formalized health care system; and working closely with and in communities, schools, and religious facilities[9, 10]. A concurrent analysis of our research showed that CHWs played a crucial role in demand generation in Nepal, Senegal, and Zambia[11–14]. The WHO health policy guidelines for CHWs program (2018) recommends the following: 1) community engagement in selection, 2) training, 3) remuneration, and 4) supervision. Coupled with other strategies, this policy aims to increase the efficiency and productivity of CHWs towards service delivery[8]. Yet, despite these recommendations, frontline health workers in many countries – the vast majority of which are women - remain unpaid and are provided minimal training and supervision[15]. Evidence suggests that scaling up and sustaining CHW programs requires effective program design and management, including adequate training, supervision, motivation and funding, community acceptability, and support from political leaders and other healthcare providers[16].

The purpose of this analysis was to identify factors of volunteer community health worker programming that successfully contributed to vaccination initiatives in Nepal, Senegal, and Zambia. We describe 1) CHW programming organization-cadres, roles and responsibilities, 2) CHW motivation-recognition, tangible incentives, capacity building, empathy and compassion; and, 3) trust-positive social capital, mutual trust-through community engagement and knowledge sharing. We discuss how these factors have contributed to success in service delivery and increased routine immunization coverage in the context of the three countries. Program implementation strategies involving active engagement of CHWs in Nepal, Senegal, and Zambia may offer insight into what are critical contributors to immunization access and intent to vaccination.

## Methods

We applied a multiple case study design to explore critical success factors of vaccine delivery systems in three countries with high vaccination rates: Nepal, Senegal, and Zambia[17]. This analysis is nested within the Vaccine Delivery project within the Exemplars in Global Health program[17, 18]. We employed a qualitative analysis to investigate factors that contributed to high and sustained vaccination coverage through key informant interviews (KIIs) and focus group discussions (FGDs) at the national, regional, district, health facility, and community levels of the health system. We triangulated these findings with quantitative analyses using publicly available data.

In this analysis, we selected three themes-organization, motivation, and trust as they relate to CHWs and how these contributed to increased immunization coverage in Nepal, Senegal, and Zambia. We provide operational definitions of the themes and some specific examples of the components as they emerged from data (Table 1). The protocol of the primary analysis, along with individual case studies of countries and a synthesis case study, are published separately[12–14, 17].

**Table 1.** Definitions of themes as derived from data.

| Domain | Definition and Examples |
| --- | --- |
| 1. Organization | Expansion of CHWs with different cadres, and roles and responsibilities; may be based on the scope of work, reimbursement, qualifications, language, education level, geographic region, and their distinct roles and responsibilities. These CHWs work together to improve community health and increase vaccination rates. |
| 1.A. Cadre | Specialized types of CHW providing immunization-related services in the communities<br>• Female Community Health Volunteers, Mother groups, Village Development Committees (Nepal)<br>• *Agents de Santé Communautaire, Matrones, Dispensateur de Soins à Domicile, Bajenu Gox, Relais* (Senegal)<br>• Neighborhood Health Committees, Safe Motherhood |
|  | Action Groups, Growth<br>Monitoring Promoters (Zambia) |
| 1.B. Roles and responsibilities | Work and functions of CHWs related to immunizations activities, based on specific age ranges of the CHWs, ethnicities, language skills and qualifications, including:<br>• Defaulter tracing which involves finding children who missed their scheduled immunization.<br>• Outreach planning<br>• Data collection and reporting<br>• Community mobilization<br>• Public awareness and education |
| 2. Motivation | The reason, desire (e.g., empathy, compassion), or willingness of a volunteer community health worker to do their work [19-21]. |
| 2.A. Recognition | Identification of CHWs by the communities and healthcare workers as integral members of healthcare team due to their knowledge and close |
|  | understanding of the communities they serve. Examples include:<br>• Respect by community members<br>• Support from community<br>• Value placed on the work of CHWs<br>• National, regional or local celebrations |
| 2.B. Tangible incentives | Factors - direct or indirect, financial or non-financial - that increase motivation to engage and perform well, which include specific forms of reward[22].<br>Examples include:<br>• Money, including salary and remuneration for specific training<br>• Food, usually staples of diet<br>• Childcare<br>• Harvesting assistance<br>• Bicycles, uniforms, raincoats, stationary<br>• Certificates, including-training completion<br>• Free healthcare services |
| 2.C. Capacity building | Includes training and supervision of CHWs and the downward impact on the community, including:<br>• Increased self-efficacy to perform responsibilities<br>• Increased knowledge of diseases, diagnoses, treatment, and facilitation<br>• Contribution to program planning and decision-making |
| 2.D. Empathy and compassion | The ability to understand and share the feelings of others, show concern for the suffering or misfortune of others, and prioritize the needs of other community members - all of which motivated CHWs to perform their roles. |
| 3. Trust | Positive social capital or social relationship, as an element of truth telling, empowering others through knowledge. |
| 3.A. Community engagement | The involvement of communities in decision making and in planning, design, governance and delivery of services[23].<br>• Community members involvement in selection of CHWs<br>• CHW as residents of their respective communities<br>• CHWs speak the same language as community members<br>• CHWs relaying health information to community members and sharing feedback from the community to healthcare workers |
| 3.B. Knowledge sharing | Activities through which information and skills are shared between individuals or groups can include frequent interaction, social relationships between CHWs, communities, and health workers[24]. |

### 2.1. Study settings

Nepal, Senegal, and Zambia were selected based on their success at achieving high and sustained growth in early childhood vaccination, based on historical (i.e., 2000-2018, up to the point of starting this project) DTP1 and DTP3 coverage estimates. Details are explained elsewhere [17]. Three regions within each country were identified in consultation with national stakeholders and available data[12–14]. Ministry of Health (MoH) officials and in-country partner organizations facilitated site selection and data collection activities. Each country has a slightly different name for their Ministry of Health (e.g., Ministry of Health and Population in Nepal; Ministry of Health and Social Action in Senegal), but all will be referred to as “MoH” throughout for simplicity.

### 2.3. Qualitative data collection and analysis

Qualitative data were collected at the national, regional, district, health facility, and community levels. Data collection was from August 2019 to April 2021. The interview guides were informed by the Consolidated Framework for Implementation Research (CFIR) and the Context and Implementation of Complex Interventions (CICI) frameworks [25, 26]. KII and FGD guides were translated into national and regional languages by research assistants, piloted before use, and adjusted iteratively throughout data collection. An initial list of KIIs was developed with local research partners and MoH officials; snowball sampling was used to identify additional key informants. Caregivers and CHWs were recruited for FGDs from health facility catchment areas with the assistance of local health staff. The duration of KIIs and FGDs averaged one and a half hours. KIIs and FGDs were audio-recorded with the permission of participants. Research files, recordings, and transcriptions were de-identified, password protected, and kept on a secure server. The activities are summarized in Table 2.

**Table 2.**
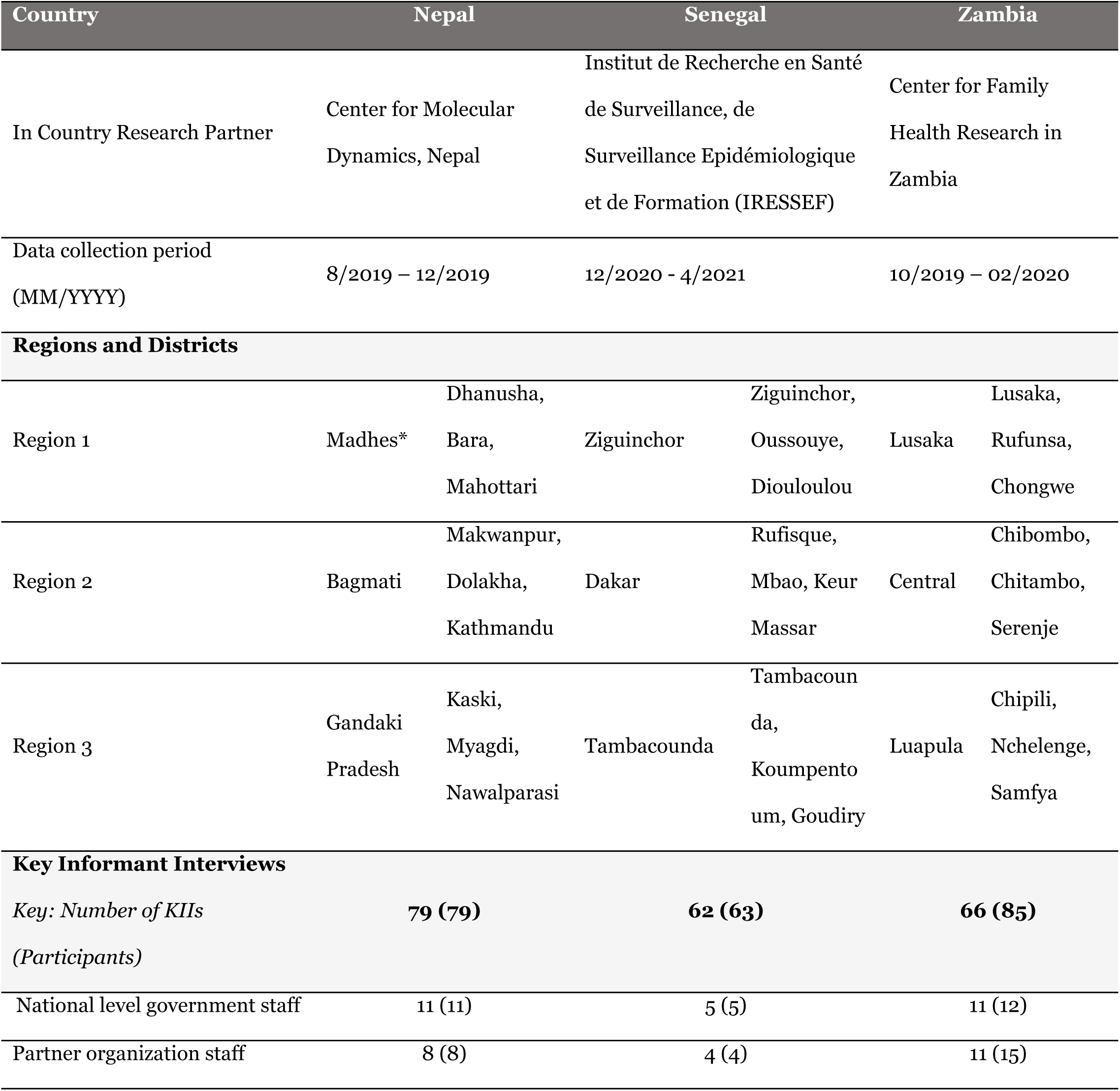
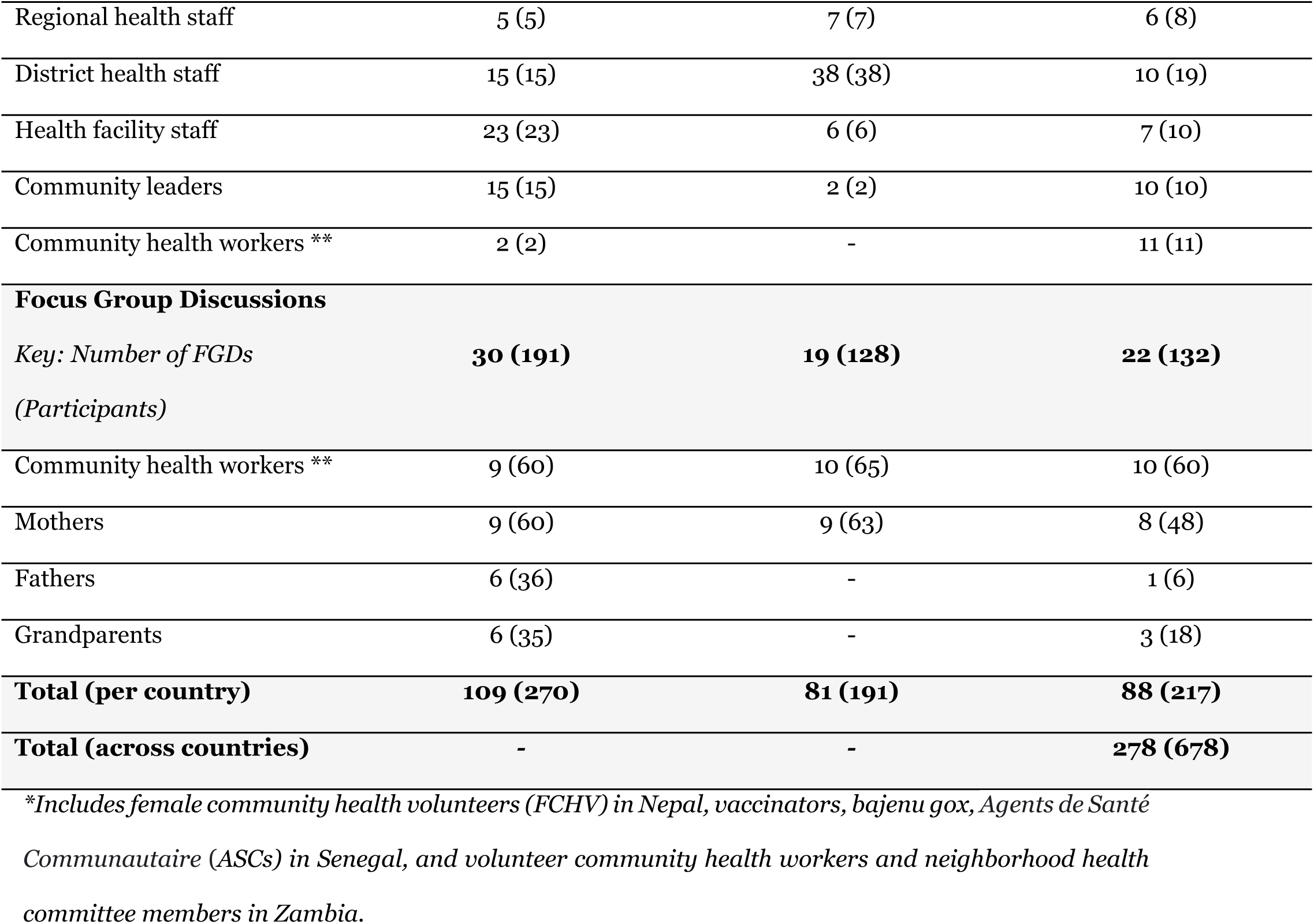
Summary of countries, partner organizations, regions, districts and data Collection Activities. [27]

### 2.4. Ethical approval

The study was considered exempt by the Institutional Review Board committee of Emory University, Atlanta, Georgia, USA (IRB00111474), and was approved by the National Health Research Committee (NHRC; Reg. no. 347/2019) in Kathmandu, Nepal; the National Ethical Committee for Health Research (CERNS; Comité National d’Ethique pour la Recherche en Santè) in Dakar, Senegal (00000174); the University of Zambia Biomedical Research Ethics Committee (Federal Assurance No. FWA00000338, REF. No. 166-2019); and the National Health Research Authority in Zambia. All participants provided informed consent, either written or with their thumbprint, in the presence of a witness.

## Findings

We analyzed strategies and delivery methods of voluntary CHWs and how they contributed to routine vaccination programming in Nepal, Senegal, and Zambia. First, we developed a framework specific to CHWs and their involvement in vaccine delivery and uptake. We included aspects of our general research framework and other scales and frameworks specific to CHWs[28, 29]. We found that CHWs’ contribution to improvement in healthcare delivery and utilization in different settings aligned with the broad domains around cultural congruence, social support, mutual trust, and others[20, 28, 29]. We then focused on the themes of organization, trust, and motivation (Fig. 1) to compare success factors across these countries.

**Fig. 1.**
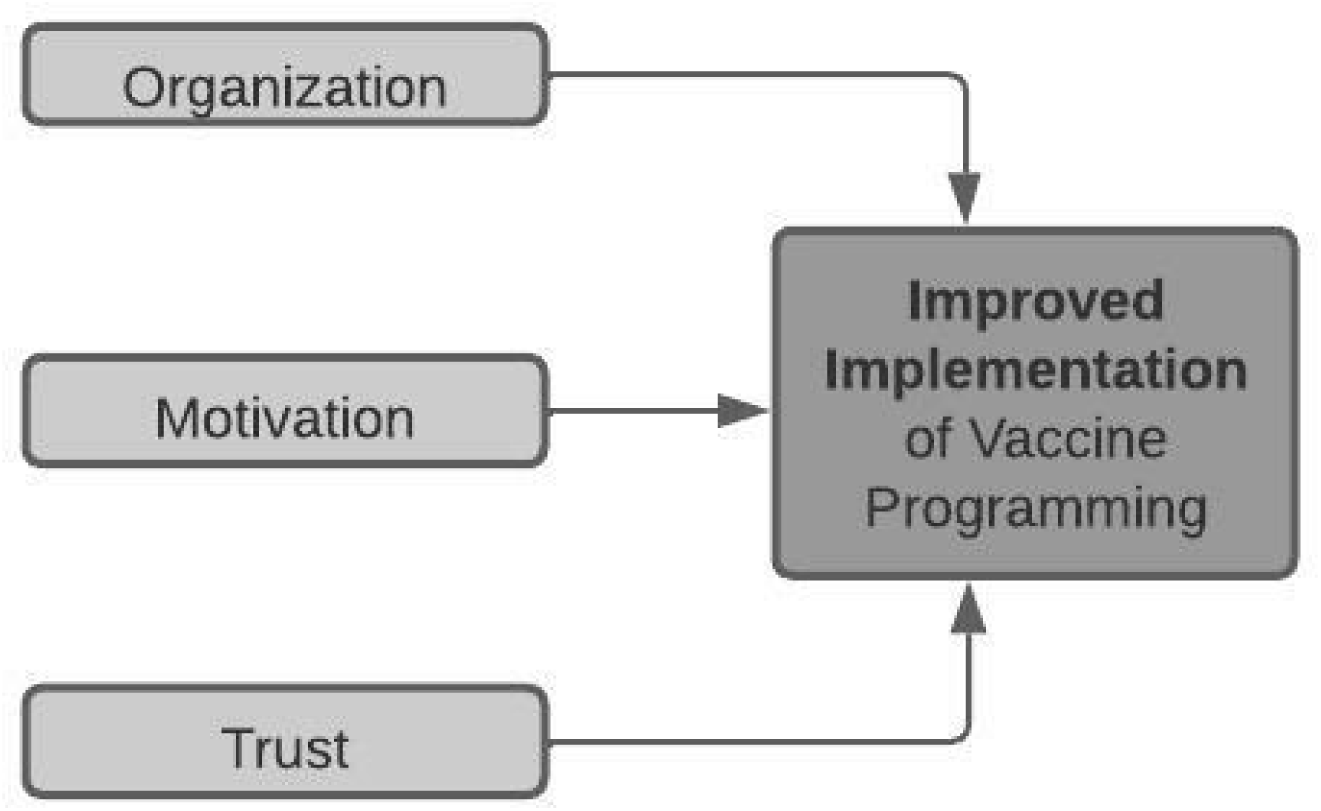
Summary figure of CHW domains that shaped vaccination in Nepal, Senegal, and Zambia.

For this analysis, we chose to focus on the roles of volunteer CHWs, recognizing that in all countries, at least one of the cadres (e.g., *bajenu gox*) was more trusted by communities and was instrumental in the vaccination programming (Table 3). CHWs capitalized on their strong community role to educate community members, combat misinformation, conduct outreach and provide personalized service to caregivers. Themes around 1) organization, 2) motivation, and 3) trust were derived from the data and contributed to improvements in vaccination program implementation (Table 1). Organization through expansion of CHW types motivated CHWs to carry out their roles and responsibilities related to vaccination. Trust that communities and healthcare workers expressed to CHWs through respect and value placed on work contributed to CHWs’ motivation resulting in the improved implementation of vaccination programming.

**Table 3:**
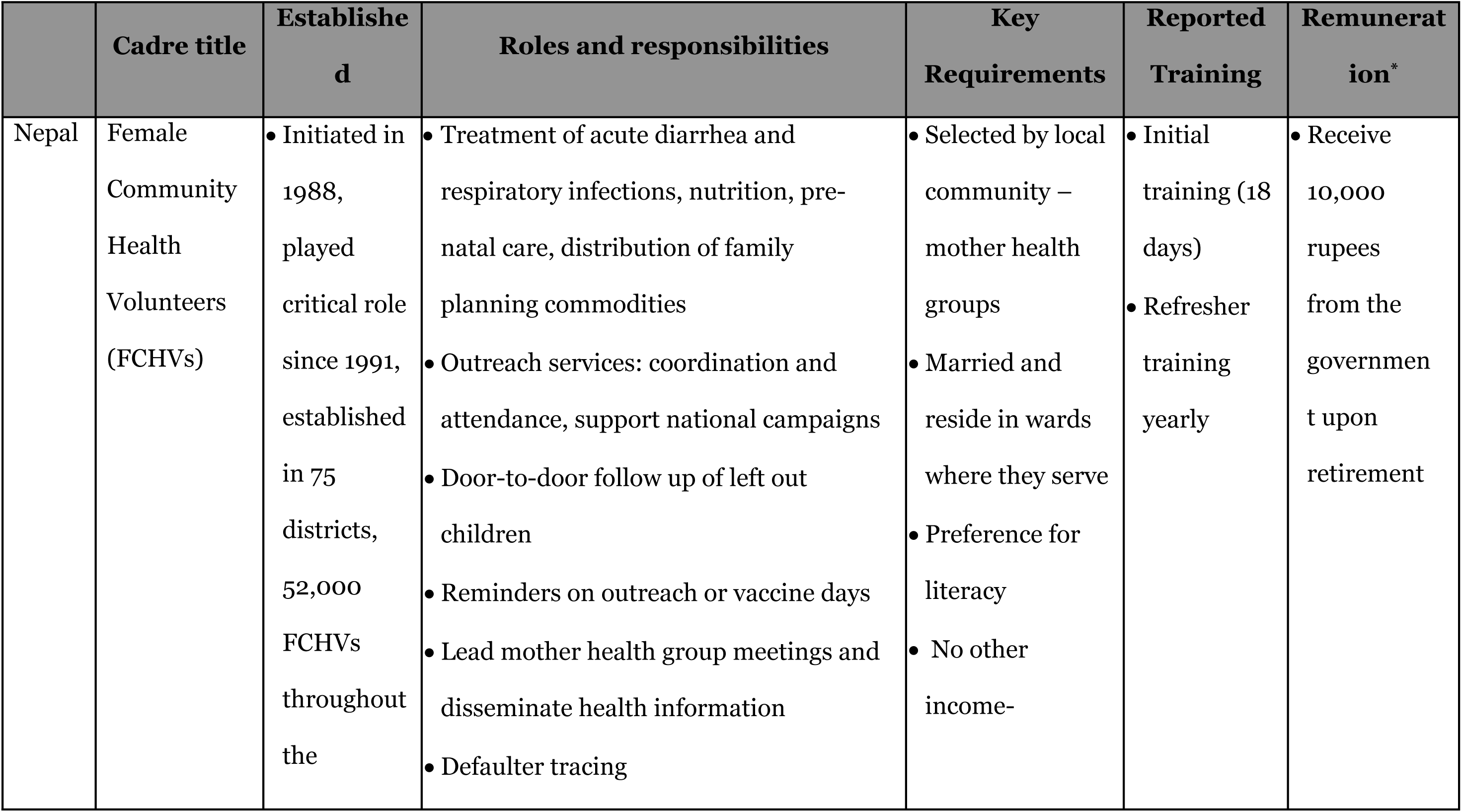
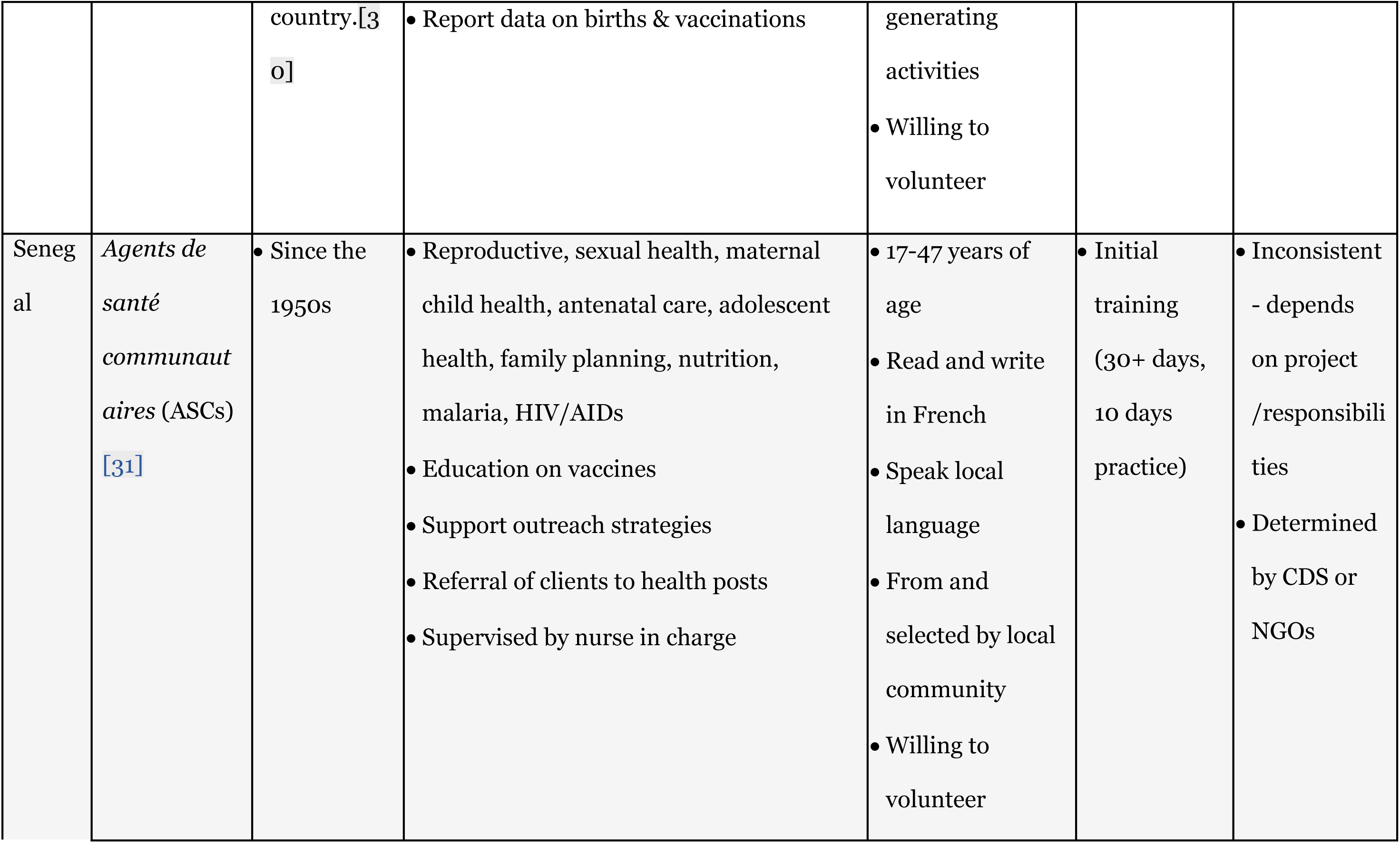
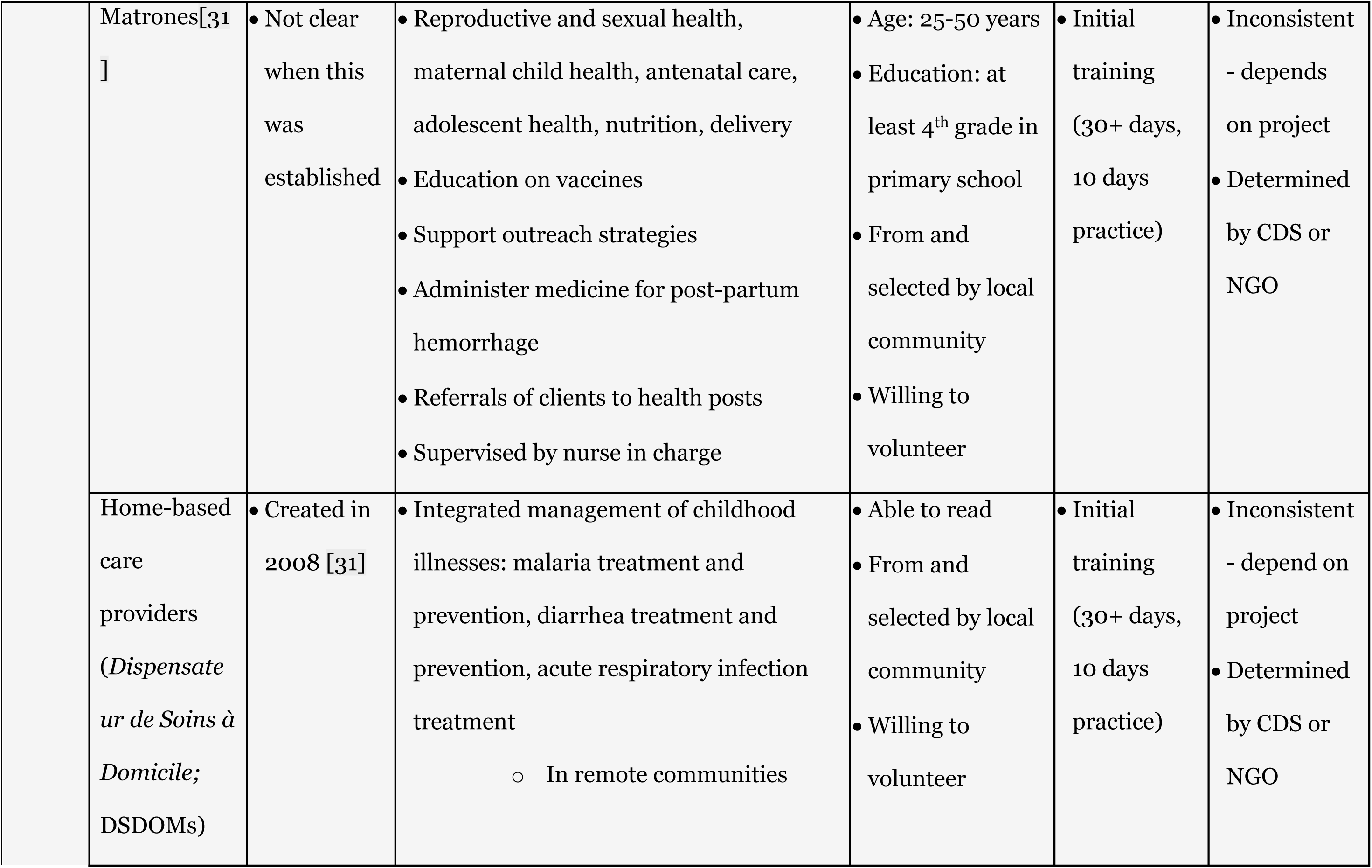
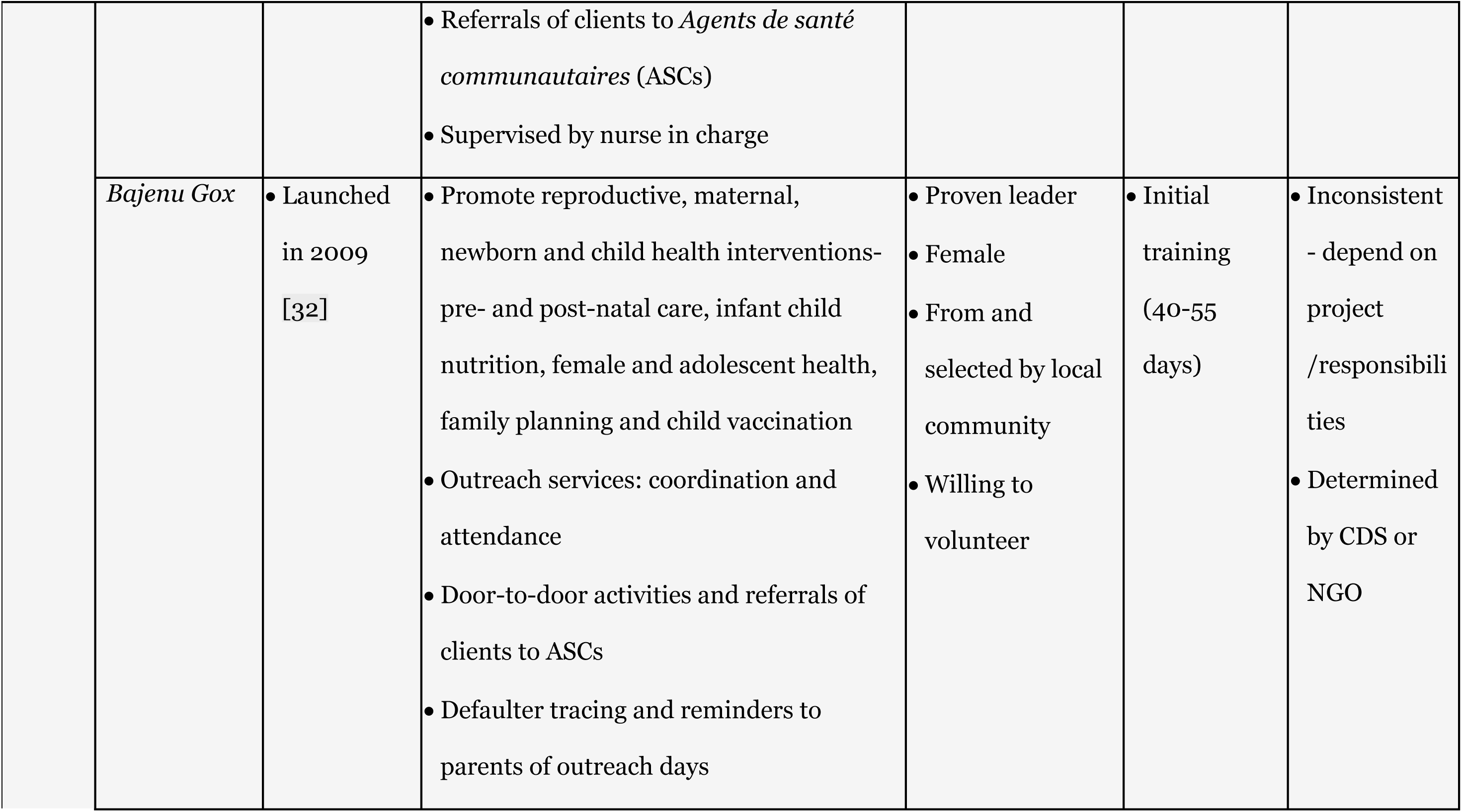
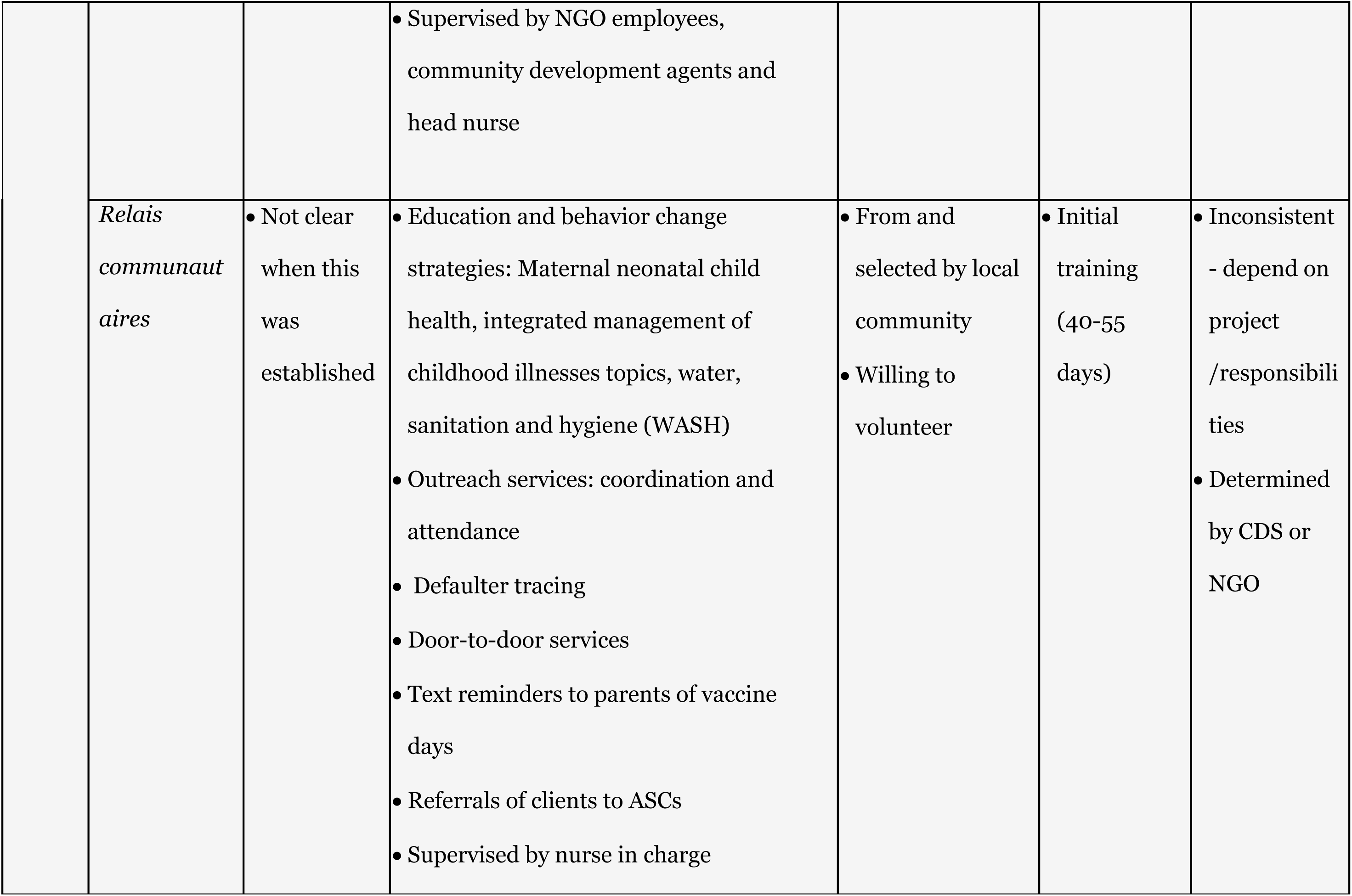
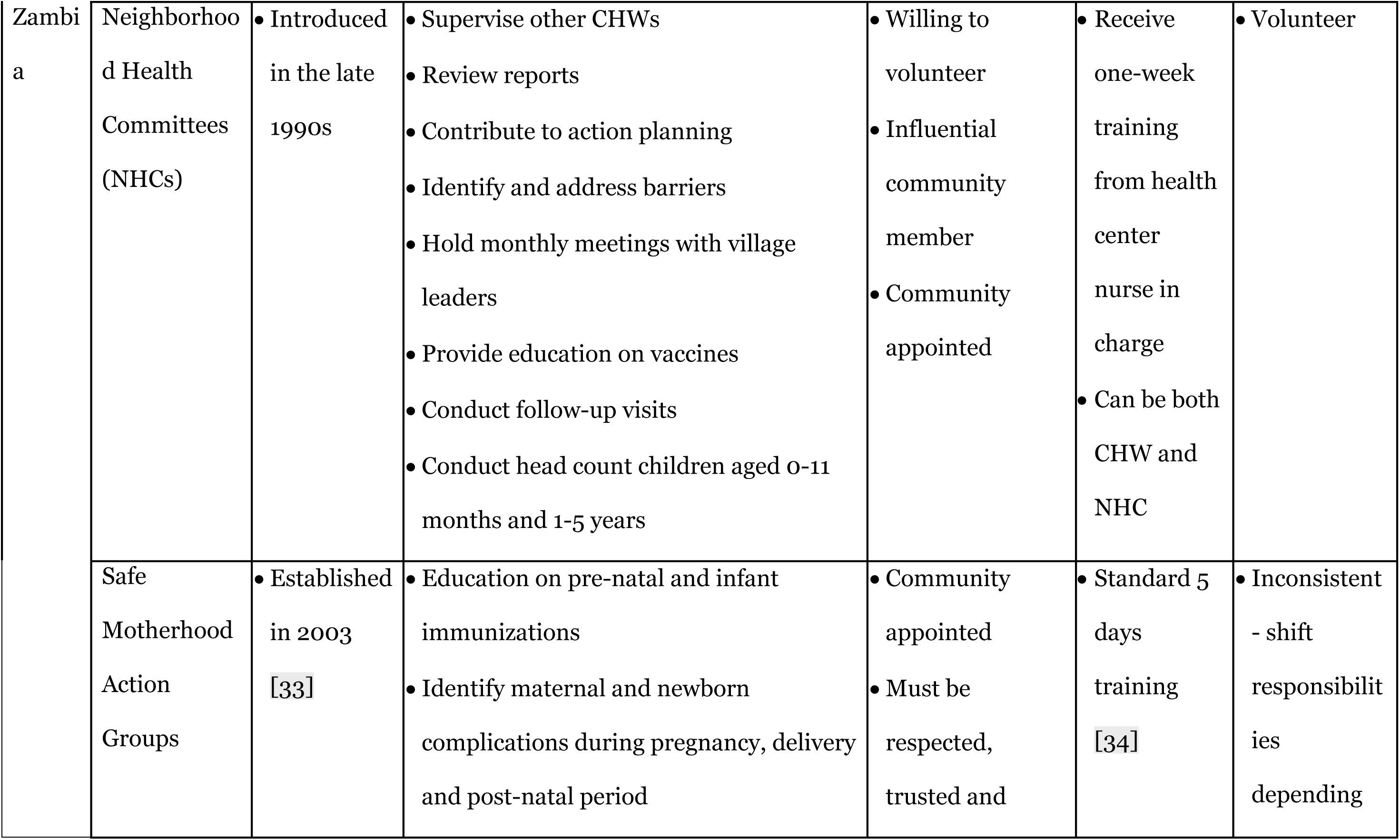
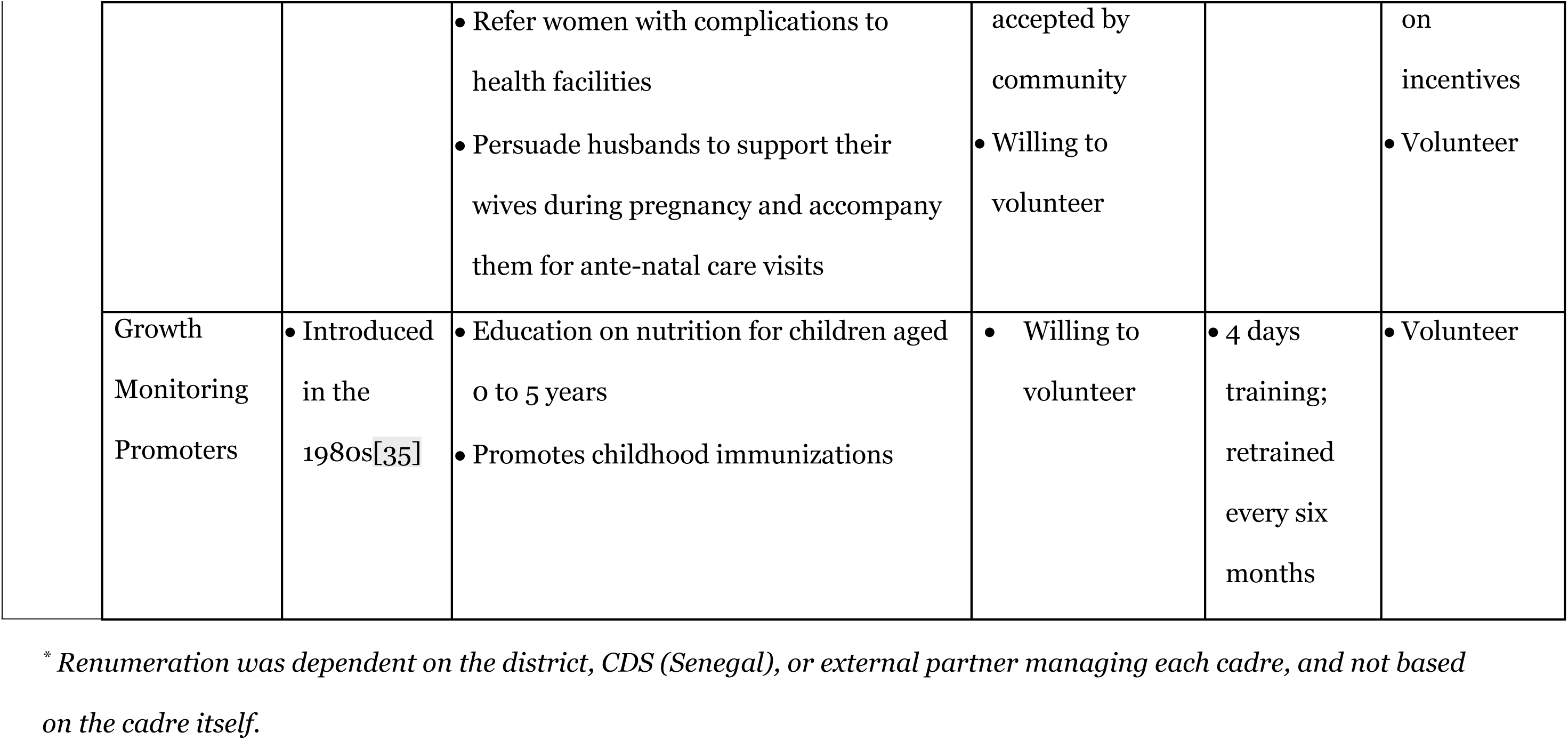
Community Health Workers cadres, roles, recruitment requirements, training and remuneration by Country.

### 3.1. Organization

In all three countries, key informants reported that recently introduced cadres and diversification of roles and responsibilities improved implementation of vaccination programming. The roles and responsibilities of CHWs shifted and often expanded, and this was subsequent with increased funding and focus on health services strengthening. Governments created more positions/cadres, defined, and aligned responsibilities with specific qualifications, which bolstered frontline health outreach and services (Table 3). Current volunteer CHW’s roles across the countries required specific qualifications, training, and responsibilities. CHW roles included generating public awareness, dispelling misinformation, defaulter tracking, and data collection.

#### 3.1.1 Cadre

Each country had different types of CHWs who played specific roles in immunization programming. These cadres were created to distribute the heavy workload, assign specific roles and responsibilities to specialized staff, and improve overall coordination and delivery of services. In Nepal, FCHVs played a major role in immunization, they created awareness on immunization, and linked the communities to the healthcare facilities. Senegal had five different cadres (Table 3) involved in different aspects of immunization. In Zambia, neighborhood health committees, safe motherhood action groups and growth monitoring promoters played a critical role in improving vaccination coverage through the roles and responsibilities they held as discussed in the next section.

The specific characteristics and detailed elaboration of each cadre of the CHWs in Nepal, Senegal, and Zambia are outlined in Table 3.

#### 3.1.2 Roles and Responsibilities

In the three countries, at least one CHW cadre exists. In Senegal and Zambia where more than one CHW cadre existed, most of CHWs performed similar roles. The lack of defined roles and localized training led to overlapping or deficient coverage of health services by cadres in Zambia and Senegal.

In Nepal, while vaccinations are carried out by salaried health workers, key informants indicated that FCHVs, “the backbone of the health system,” receive the most credit nationwide for improved immunization coverage. With the increase of paid frontline health workers in 2010, the responsibilities of FCHVs shifted to building awareness of available health services and their benefits at different community levels, including local mothers’ groups (Table 3).

> *“FCHVs and the mothers’ group provide information regarding immunization campaigns or [the] regular immunization program. For new vaccines, it needs to [be] discussed with FCHVs and [the] mothers’ group - unless there is support and involvement in the program, it will not be successful because they are the one who build awareness and give knowledge and information to the people in the grass root level.” (Ministry, National level, Nepal)*

In Senegal, *relais* and *bajenu gox* conduct more targeted vaccine outreach although the five cadres of CHWs serve complementary roles in delivering frontline health services (Table 3). KIIs and the literature identified the *relais* and the *bajenu gox* as critical to the success of the vaccination program, and mothers referred to them as “real fighters” for the health of children. Community promotion actors are mobile, focusing on visiting communities, neighborhood organizations, and homes to promote awareness of and create demand for disease prevention and health promotion activities, including vaccinations. Community members lauded the mobilization and the motivation these actors inspired which contributed to the improvement of vaccination programming.

> *“The involvement of community actors helps them a lot because the community actors live with the populations and know the families. Today, even after the counting, they cannot determine the house of someone or they are given the wrong telephone number. But with the involvement of community actors, they go into the neighborhoods to bring back the children.” (Community health worker, community level, Senegal)*

In Zambia, key informants discussed the Neighborhood Health Committees (NHCs) extensively. NHCs are community-appointed volunteers whose primary role is to coordinate and manage other CHWs on behalf of health facility staff, including CHW recruitment, training, and attrition. NHCs also ensure that CHWs accurately report monthly activities and represent both CHWs and community members in annual action planning. NHCs’ responsibilities have increased in the last two decades to accommodate the increasing number of CHW roles across multiple health sectors, bridge the gap in human resources, and institutionalize the link between communities and the health system. Many individuals who hold NHCs role also hold another CHW position so as to receive training and other incentives through disease-specific initiatives.

> *“A long time ago, as I said, we [community members] used to come in numbers here at the [health] center. It was difficult because we [the community] did not have enough [health facility] staff, so that’s how this [the NHC] program started. And they [NHCs] started talking to people so that we [NHCs] help each other because members of staff were few. We [NHCs] are grateful they trained us, and we are working.” (NHC member, community level, Zambia)*

The NHCs in Zambia were involved in action planning, identifying annual priorities, setting objectives, selecting strategies, determining indicators to monitor and evaluate activities, and estimating required activity resources and budget. Through their involvement, the NHCs indicated ownership over these responsibilities, which were formalized in the Reaching Every District (RED) program manuals.

### 3.2. Motivation

Community health workers in all three countries emphasized the meaning and value they found in their work. This was through recognition of their effort, incentives, and knowledge gained through training. CHWs in all the countries received training, which increased their knowledge on vaccinations and the present vaccine challenges. However, training varied and was sometimes inconsistent within specific countries. Supervision was done to ensure that CHWs performed their roles well; though, inconsistency in supervision was experienced in all countries, with key informants indicating workload and insufficient human resources as the barriers. Beyond the individual level, CHWs were motivated through the declaration and celebration of their villages by the national government as fully immunized, and successfully reaching all children in their villages through the implementation of Reaching Every Child (REC)/Reaching Every District (RED) strategies in Nepal, Senegal and Zambia, respectively.

#### 3.2.1. Recognition

In all three countries, CHWs stated that recognition for their contributions and their status in the community were motivating factors. Communities, districts, and regions recognized the efforts and successes of CHWs through ceremonies, certificates, and media recognition. Community selection showed recognition and respect for the individual chosen to be a CHW. Particularly for women, CHWs gained stature through imparting valued knowledge, supporting caregivers, and contributing to positive health outcomes.

> *“Our motivation is we have respect through this work. Everyone praises our work. They praise our contribution towards work for pregnant women, children and delivery of pregnant women.” (FCHV, Nepal)*

In Nepal, FCHVs appreciated the recognition for their role in increasing vaccine coverage. Some FCHVs received certificates. Villages, districts and provinces were declared fully immunized and celebrated by the national government through the Full Immunization Declaration (FID) Program in a bottom-up approach, and this motivated community workers and volunteers, and facilitated community buy in to immunization. Children who are fully vaccinated receive an official vaccination card with complete information on vaccines received from their health post for documentation. Increased public awareness and FCHV activities support the success of the Full Immunization Program (FIP) – which promotes urgency and action among all key players, including caregivers, religious and community leaders, and teachers.

***“****We are all working together so that the children won’t miss vaccines. We can assure you that every child in our area have got vaccines.” (FCHV, Nepal)*

#### 3.2.2. Incentives

In all countries, incentives were crucial to engage and motivate CHW volunteers. Though inconsistently distributed, tangible extrinsic resources - including money, food, bicycles, uniforms, stationary, childcare, and harvesting assistance - motivated CHWs and could reduce any intra-household tensions caused by absences due to the program. When CHWs areas of jurisdiction reported low vaccination coverage, especially in comparison to other districts or regions, their motivation to improve outcomes increased.

> *“We do recognize them [FCHVs]. We do a public audit and take decisions on whom we should give credit; that is recognized by the community and system so that they become more motivated.” (Regional Director, Nepal)*

Intrinsic incentives were reported as critical motivators, particularly for those in Nepal and Senegal. CHWs highly respected their communities, and they served voluntarily because of the mutual respect between community and CHW. Seeing children in their communities protected from diseases and growing up healthy was additional motivation. FCHVs had pride and expressed happiness if all the children within their catchment areas were fully vaccinated. A FCHV in Nepal stated that “they worked for the good of the community.”

However, even with efforts to recognize CHW contributions, CHWs in all countries felt underappreciated by the national government. They expressed that the appreciation they received was more evident in the community than by the government. In all countries, CHWs felt they lacked commensurate compensation for their labor; they were required to commit to being a volunteer. Retention suffered because in some instances, CHWs were forbidden to engage in income-generating activities, and compensation was often inconsistent. CHWs families often expected them to complete household labor even though they were engaged in volunteer responsibilities. Community health volunteers often supported and contributed to the work of paid CHWs and head nurses, resulting in additional workloads without appropriate compensation. Additionally, countries’ projections for labor were unmet, leading to heavy responsibilities for CHWs.

#### 3.2.3. Capacity building

CHW’s knowledge increased through training and supportive supervision; in many cases, the opportunity to learn new skills and knowledge promoted self-value and enhanced their capacity to support the communities in the three countries. CHWs received formalized training through national health programs, NGOs, and multi-lateral partners; however, the length, breadth, and depth of training differed by country and trainer (Table 3). Key informants valued NGO and multilateral training, which provided financial incentives. Some FCHVs and *bajenu gox* reported the benefits of CHW literacy promoting groups. In all countries, CHW utilized their community knowledge to contribute to decision-making, which ultimately contributed to planning at higher levels. CHWs had frequent interactions with the local population, as they resided in those communities. This facilitated their deeper understanding of the community’s needs, which they escalated during planning meetings. The frequent interactions between CHWs and community members enhanced community empowerment, enabling development and maintenance of social norms related to vaccination.

Supervision of CHWs occurred in all countries. Supportive supervision increased CHWs self-confidence through cross-learning and skills sharing facilitated teamwork, and enhanced relationships with healthcare workers. In Senegal, *bajenu gox* received supportive supervision while working in communities. Their supervisors recorded field visit results on a standardized checklist, and sent supervision reports to the district medical offices for review. However, supervision in the field may be rare due to a lack of human resources. In Zambia, the NHCs have a supervisory position over CHW reporting, and play an essential role in data collection and evidence-based decision-making. NHCs were motivated by these responsibilities because they elevated the NHC’s position, and described the emphasis on community-driven strategies to combat health challenges. Health facility staff and supervisors from the districts include NHCs in meetings which provide encouragement. According to national policy, by encouraging the possession and responsibility of data, the frontline workers review their performance against their stated goal, identify gaps that exist, and discuss ways to improve vaccination coverage.

> *“There is the NHC that spearheads all community activities because all the CBVs, all community volunteers, they report through the NHC.” (District Nursing Officer, Zambia)*

Although CHWs in all countries desired consistent and increased opportunities for training; these differed significantly depending on the location and human, financial, and material resources.

#### 3.2.4 Empathy and compassion

CHWs expressed being motivated by their sense of empathy and compassion for their communities in all three countries. CHWs understood and shared the feelings of their fellow community members and showed concern for their suffering and misfortune. This motivated CHWs to perform their roles well since they felt invested in the health of the community at large in addition to children’s health. CHWs understand the lived experiences of the community members and have compassion for the potential morbidity and mortality of children when not immunized against vaccine-preventable diseases. This increased their effort to focus on improving the health and wellbeing of children in their communities.

> *“Vaccine prevent from many diseases that is also why people bring their children for vaccination. There were more Pneumonia cases and diarrheal diseases before but nowadays there are very less children who suffer from Pneumonia and Diarrhea. If we hear that some children have suffered from diseases then we feel very worried. We also feel responsible towards those children.” (FCHV, Nepal)*

### 3.3. Trust

CHWs increased community members’ trust in the formalized health system. CHWs were respected members of the community, with some individuals regarded as opinion leaders; this trust was facilitated by the selection of CHWs from the communities in which they lived. CHWs connected with the community by promoting and prompting vaccination. In Senegal and Zambia, CHWs facilitated the adaptation of REC/RED strategy to local context, ensuring community buy-in. These strategies required frequent interaction with community members - providing education, action planning, conducting surveys, and tracking vaccination records. Through such interactions, social relationships were built, which strengthened community trust in CHWs and the activities they carried out within these communities. Although all CHW cadres had roles they served towards improving the health and well-being of the communities, respondents identified a cadre of CHW that they most trusted and felt they had the most investment in their health and their children’s health (Table 3).

#### 3.3.1 Community engagement

Communities in all three countries were involved in decision making, planning, design, governance and delivery of services. In Nepal, FCHVs are required to work in the communities they reside in; CHWs reported that this generated trust and promoted culturally and contextually appropriate messaging to diverse stakeholders. FCHVs educate community members in mothers’ health groups (MHG), ensuring the group’s integration into their assigned communities to share health information with community members. As FCHVs were embedded in the communities where they conducted outreach, their messaging incorporated differences in religion, tribe or caste, ethnicity, and language.

> *“I: How much do community members trust the sources of Vaccination information? //P4: Yes, they have trust. //All at a time: yes, we believe on them. //P5: Because they will also learn some information from here and provides us other information. // P4: They will learn from here and teach us.” (Mothers, Nepal)*

To address the lack of necessary human resources for community health outreach and support in Senegal, the Ministry of Health, under a USAID-funded community health plan, instituted the *bajenu gox* (godmothers) program in 2009. The *bajenu gox* were trusted older women - “proven female leaders” - living in the community who capitalized on their age and social role to support pregnant women and their families in pre- to post-natal health, including immunizations (Table 3). *Bajenu gox* duties were specifically targeted at building relationships of trust with community members. Many mothers specifically praised the *bajenu gox* for taking the time to explain diseases that vaccinations targeted, when vaccinations were happening, follow-up and individualized planning.

> *“There are criteria that really push us to choose bajenu gox who are listened to, who are considered wise individuals, who have influence, who have a voice that carries when it comes to the health of the population.” (EPI Focal Point, Senegal)*

*“Special mention to our bajenu gox. We are all women, and we see the support they give us; they are our mothers and at the same time our bajenu gox. They leave all their activities to follow us and our children…each time there is a vaccination to be given; they identify the people concerned and accompany them to be vaccinated. Because vaccination is very important, before we said that without it, we could catch all kinds of diseases such as measles and others, but now we have more vaccines and thank God we have fewer diseases, and all our children are protected.” (Mothers, Senegal)*

Trust between the health workers and the *bajenu gox* was enhanced through supportive supervision. NGO employees, community development agents, and head nurses were intended to provide supportive supervision for *bajenu gox* while they were working in communities, record field visit results on a standardized checklist, and send supervision reports to the district medical offices for review. This allowed for confidence and trust with service delivery among *bajenu gox*, the communities, and health care workers.

#### 3.3.2 Knowledge sharing

In all countries, information and skills are shared between individuals or groups, and this included frequent interaction, and social relationships between CHWs, communities, and health workers. CHWs were involved in providing health education at facilities, thus creating awareness about immunization. In Nepal, health and immunization education were presented in monthly meetings; content was selected and tailored depending on literacy, misinformation, and cultural norms. In women’s savings groups (groups of women who come together to save, and lend, money to members), FCHVs emphasized the link between children’s health and future educational and occupational opportunities. FCHVs also approached caregivers individually, discussing potential barriers to immunization, addressing specific information caregivers lacked, and encouraging immunization access.

> *“FCHV sisters also help us and teach us about [health]. They also teach and train us to get vaccination for the children… We trust [the information] and it is for our children. It is better not to suffer from any diseases. First, we need to have proper information about it and only then trust it.” (Female Community Health Volunteer, Nepal)*

*“From the time of pregnancy till the birth of the child, the people trust us in this process. People trust us more than their family members and come to take suggestions from us if they face problems. Also, they do as we suggest.” (Female Community Health Volunteer, Nepal)*

In Senegal, the *bajenu gox* included multiple family members in educational outreach on vaccinations and maternal and child health. *Bajenu gox* held education groups for adolescent girls in addition to women of all ages to target misinformation and mediate between different age groups. In recent years, *Bajenu gox* have also conducted outreach with fathers’ groups, aiding in full family approval of childhood vaccinations. In Zambia, NHCs played a prominent role in community sensitization, outreach, and vaccine education in most of our research communities in Zambia. NHCs were a trusted source of information for community members since they were recruited by community leaders and worked closely with health facility staff to coordinate outreach sessions. Consistent follow-through of outreach sessions contributed to community members’ trust in NHCs and, by extension, the health system. NHCs also worked to dispel myths and misconceptions about vaccinations by educating parents on the benefits. For example, if the barrier to access were due to a parent’s competing priorities, the NHC member would escort the child to the outreach event.

> *“There is some time on the radio, and they call the NHC to come and talk about health issues. They encourage, especially in the village. They announce that next week, ‘do not delay the children to come for vaccinations.’ Yes, because of the community radio, they let anyone talk. Yes, especially the NHC, they talk a lot on encouraging other vaccines.” (Head Woman, Zambia)*

CHWs were involved in ensuring an accurate community registry (organization), had knowledge of communities and children’s vaccination status (trust), and a drive to conduct specialized outreach to increase coverage (motivation).

## Discussion

We conducted a qualitative study to assess factors that motivated or inhibited engagement of the CHW programs in three countries between 2000 and 2019 - Nepal, Senegal and Zambia. Our findings suggest three domains behind community health workers - organization, motivation, and trust - strongly contributed to improve the implementation of health programming and may have positively influenced vaccination during our targeted time period. Organization through expansion of CHW types motivated CHWs to carry out their roles and responsibilities related to vaccination. Trust between communities, healthcare workers, and CHWs contributed to CHWs’ motivation - resulting in the improved implementation of vaccination programming. Our findings are policy relevant for other countries looking to expand vaccination through CHW platforms; however, those looking to apply these findings should be mindful that adaptation to local context is crucial.

### Organization

Distinct cadres of CHWs had specific roles and responsibilities assigned to them in the three countries, which aligned with strategies implemented in other countries[36]. Diversification of roles increased the sense of responsibility, confidence and commitment of CHWs to their work. This study, however, pointed out insufficiency in CHWs supervision, like findings from other studies within similar contexts[16, 37]. Since CHWs in middle and low-income countries are not healthcare professionals, increased supportive supervision may increase and enhance retention of knowledge, thus increasing their confidence. CHWs working together with other health workers and receiving required supervision could increase their confidence in work, and their motivation to work harder, thus improving their performance.

### Motivation

Motivation of CHWs was increased through extrinsic “work for the good of the community” and intrinsic incentives-certificates, recognition, stipends, and trust. Social cohesion-the “capacity of society to ensure wellbeing of all its members by minimizing disparities and avoiding marginalization” in regard to the health and wellbeing of children was seen in the three countries [38]. Capacity building, public awareness, involvement of CHWs and communities in different health programs, and the recognition of childhood vaccination as a human right enhanced a generational shift in existing norms - hence communities increasingly embracing vaccination. In Asia and Africa, a multiple case study found that extrinsic motivation to improved CHW work, similar to the findings of this study. Organization and cultural context shaped the motivation of CHWs, as individual beliefs, pleasure and happiness drawn from wellbeing of children in the communities was also motivating [39]. In Zambia, safe motherhood action groups expressed happiness with their work because they helped saved women’s lives [40]. In Nepal, Senegal and Zambia, CHWs were part of the communities they served, increasing their sense of belonging and trust, therefore increasing their motivation to dispense their duties.

CHWs are often the backbone of the frontline health systems in many parts of the world. Yet, this workforce, most of whom are women, remain unpaid, often resulting in undue psychosocial stress, food insecurity, and time burdens[41, 42]. Unpaid work, including the roles of female CHWs, leave CHWs time poor, being unable to meet their basic needs, or participate in social or political activities - contributing to the widening equity gap and expanding the poverty cycle[43]. CHWs’ great contributions to healthcare delivery is evident; however, limited attention has been placed on their remuneration. Nevertheless, payment of living wages to CHWs is a fundamental right[44]. WHO now recommends remunerating practicing CHWs for their work with financial package commensurate with job demands, complexity, number of hours, training and roles and responsibilities[8]. Nepal, Senegal and Zambia provide incentives to CHWs; however, this is incommensurate with their work[22]. Similarly, countries like Sierra Leone and Liberia have policies that guide the remuneration of CHWs, but CHWs from these countries expressed dissatisfaction with remunerations in place, highlighting the need to consider contextual factors and roles and responsibilities when planning and budgeting for CHW remuneration[45]. Although consistency in the incentivization of FCHVs was observed due to integration, and the financing of CHW programming at the national level in Nepal, Senegal and Zambia reported inconsistencies. In low- and lower-middle-income countries, CHWs are not adequately remunerated, even with long service to the community and numerous contributions to elimination of diseases, reducing the menace of diseases, and increasing the reach of immunization programmes. Furthermore, the roles of CHWs are deemed to fit into the other domestic activities, which are mostly done by women [46].

Nevertheless, the incentivization of CHWs in all countries was associated with higher motivation to create public awareness and conduct community mobilization. Countries with clear financial management systems that include budgets for CHWs have seen great contributions of CHWs in general healthcare service delivery. Ethiopia and, Bangladesh, among others, successfully implemented a CHW strategy which has seen their vaccination coverage increase [36]. However, CHWs remuneration varies between countries, because of limited political will, targeted funding, and historical underpinnings and addressing these challenges may improve CHW performance and retention - thus sustainability - of CHW programming[22, 47]. Lack of consistent income could fail to overcome intra-household pressures, address competing priorities at the household level, and ultimately lead to decreased morale and retention of CHWs. Standardizing CHWs compensation within different settings may be of value, which could be done through governments setting aside a funding stream dedicated to CHWs and considering other sources of funding to supplement existing sources. These strategies may reduce the financial challenges experienced by the CHWs within these countries.

### Trust

Trust and community ownership were generated through the CHWs selection process, which increased their motivation to provide immunization-related services to the communities in Nepal, Senegal and Zambia. CHWs shared information on vaccination in local languages and reached hard-to-reach areas and minority communities. Being dependable, accountable, and respected members of the community, CHWs understand the contexts of the communities they serve [16, 48]. This generates trust and enhances accountability through 1) CHW closeness to the community and local healthcare facilities, 2) uptake of information provided, and 3) ease in community entry, and that of the health workers thus reducing child vaccination gaps especially among the illiterate mothers[49]. Trust between caregivers and CHWs resulted in adequate knowledge and compliance with vaccination recommendations.

Changes in healthcare, including introduction of new vaccines and emerging epidemics, add another layer of work to CHWs and can be addressed through different cadres with specific responsibilities. The recent COVID-19 pandemic highlights the contribution of CHWs in creating demand for vaccination through addressing vaccine hesitancy, leveraging on their existing trust within communities [50]. Strategic approaches that allow for expanding CHW cadres are necessary to bridge the gap in human resources, and provide vaccination services to disadvantaged populations [51].

Governments have committed to addressing some of the issues that CHWs have voiced; to combat shortages of community health workers in Zambia, the central government committed to increasing the number of community health workers, investing in training, and retaining more workers. The Zambia National Health Strategic Plans and the 2010 and 2019-2021 National Community Health Worker strategies instituted guidelines to formalize and harmonize the roles of frontline workers to decrease overlap in responsibilities, as well as uniform incentives. The government committed to bringing 30,000 more workers on board and increasing specialization of cadres and workers. In 2018, Senegal’s government committed to creating more formalized training for different cadres. The 2019-2021 CHW Strategy identified disharmonized incentives as a challenge and allocated the district health office to coordinate and oversee CHW incentives (such as lunch and transport reimbursement), supplies (like protective clothing), orientation and training, and involvement in national campaigns. In Nepal, FIP worked well to motivate service providers, community workers, volunteers, and parents.

### Limitations

This study has several limitations. First, the research tools focused on the factors that drove catalytic change and did not focus on interventions or policies that were unsuccessful. Second, using qualitative methods to understand historical events was challenging; interviewees often spoke about current experiences rather than discussing historical factors. However, research assistants probed respondents to reflect on longitudinal changes in the immunization program. Third, the scope of our study was extensive, and CHWs were only one component of our research. Fourth, some policy documents – including national-level strategic plans - were not available for our review. And finally, the COVID-19 pandemic impacted data collection in Senegal, with a focus on response and safety taking priority over data collection.

## Conclusion

This study highlighted essential qualities of community health worker programs in countries with high vaccination coverage – namely, organization, motivation, and trust. Each country had distinct CHW cadres involved in the implementation of vaccination programming. Additionally, CHWs were selected from their communities - which generated trust among community members and healthcare staff. CHWs bridge the equity gap in access to vaccination services through involvement of female volunteers, and CHWs enabled wider reach of vaccination services to minority populations and populations in hard-to-reach areas who otherwise could not be easily reached by healthcare workers. Although improvements in vaccination programming were seen in all three countries, CHWs faced challenges in providing adequate services in their communities. Workload, low and inconsistent compensation, inconsistency in training duration and scope, and supervision resulted in demotivation and high turnover of CHWs in all countries. While plans for changes in the CHW programming report increased duties and responsibilities for CHWs, the governments in all countries still need to adequately address issues related to the recognition of CHWs, especially their compensation. Based on their contexts, countries should focus more on ways of addressing such challenges, and country-specific research on strategies to ensure consistent funding for CHWs would be beneficial.

## Data Availability

All data are available from the Open Science Framework website (https://osf.io/7ys4a/?view_only=739ca7a72f9749118b4aa3d2f7b655d9).

## Acknowledgements

We thank Center for Molecular Dynamics, Nepal (CMDN), the Institut de Recherche en Santé de Surveillance Epidemiologique et de Formation (IRESSEF) in Dakar, Senegal and the Center for Family Health Research in Zambia for their partnership in this study. We gratefully acknowledge the participants who gave their time and insights to help us better understand the vaccine delivery system in Nepal, Senegal, and Zambia, along with facilitators from the Ministry of Health and Population, Department of Health Services, and the Family Welfare Division of Nepal, Ministry of Health and Social Action in Senegal, and the Ministry of Health in Zambia for supporting this research. We thank Chin-En (King) Ai, Lauren Harper, Allison Wray from Emory University and Saad Omer from Yale University for their contribution to this project. In addition, we thank Sarah Chesemore, Anna Rapp, Tove Ryman, and Ethan Wong from the Bill and Melinda Gates Foundation; Kate Buellesbach, Nancy Fullman, Nathaniel Gerthe, Gloria Ikilezi, Caitlyn Mason, David Phillips, and Oliver Rothschild, Jordan-Tate Thomas, and Angela Wang from Gates Ventures for technical support; and the Vaccine Exemplars Research Advisory Group for their insights, specifically Agnes Binagwaho, Laura Craw, Carolina Danovaro, Anuradha Gupta, Heidi Larson, Penelope Masumbu, Kate O’Brien, Helen Rees, Lora Shimp, and Aaron Wallace. 

## Notes

### Competing Interest Statement

The authors have declared no competing interest.

### Funding Statement

This work was supported by the Bill & Melinda Gates Foundation, Seattle, WA (OPP1195041) with a planning grant from Gates Ventures, LLC, Kirkland, WA. MCF and RAB received the award. No commercial companies funded the study nor the authors. Bill & Melinda Gates Foundation is a non-profit and Gates Ventures is a venture capital firm. None of the authors received salaries from commercial companies. URLs to sponsors' websites is https://www.gatesfoundation.org/. Bill &Melinda Gates Foundation had no explicit say in the design, data collection, analysis, decision to publish or preparation of the manuscript. However, BMGF and GV were engaged in the project process, provided context, when necessary, but all research activities and publications are solely those of the authors.

### Author Declarations

The study was considered exempt by the Institutional Review Board committee of Emory University, Atlanta, Georgia, USA (IRB00111474), and was approved by the National Health Research Committee (NHRC Reg. no. 347/2019) in Kathmandu, Nepal the National Ethical Committee for Health Research (CERNS Comité National d’Ethique pour la Recherche en Santè) in Dakar, Senegal (00000174) the University of Zambia Biomedical Research Ethics Committee (Federal Assurance No. FWA00000338, REF. No. 166-2019) and the National Health Research Authority in Zambia.

## References

1. Orenstein WA, Ahmed R. Simply put: Vaccination saves lives. National Acad Sciences; 2017. p. 4031–3.

2. Rodrigues CM, Plotkin SA. Impact of vaccines; health, economic and social perspectives. Frontiers in microbiology. 2020;11:1526.

3. WHO. Immunization Agenda 2030: A Global Strategy To Leave No One Behind. 2020.

4. WHO. Global strategy on human resources for health: workforce 2030. 2016.

5. WHO. Health workforce 2021 [cited 2022.

6. Love MB, Gardner K, Legion V. Community health workers: who they are and what they do. Health Education & Behavior. 1997;24(4):510–22.

7. Masis L, Gichaga A, Zerayacob T, Lu C, Perry HB. Community health workers at the dawn of a new era: 4. Programme financing. Health Research Policy and Systems. 2021;19(3):1–17.

8. WHO. WHO guideline on health policy and system support to optimize community health worker programmes: World Health Organization; 2018.

9. Pérez LM, Martinez J. Community health workers: social justice and policy advocates for community health and well-being. American journal of public health. 2008;98(1):11–4.

10. Patel AR, Nowalk MP. Expanding immunization coverage in rural India: A review of evidence for the role of community health workers. Vaccine. 2010;28(3):604–13.

11. Hester KA, Sakas Z, Ogutu EA, Dixit S, Ellis AS, Yang C, et al. Critical interventions for demand generation in Zambia, Nepal, and Senegal with regards to the 5C psychological antecedents of vaccination. medRxiv. 2022.

12. Hester KA, Sakas Z, Ellis AS, Bose AS, Darwar R, Gautam J, et al. Critical success factors for high routine immunization performance: A case study of Nepal. Vaccine: X. 2022;12:100214.

13. Sakas Z, Hester KA, Rodriguez K, Diatta SA, Ellis AS, Gueye DM, et al. Critical success factors for high routine immunization performance: A case study of Senegal. medRxiv. 2022:2022.01.25.22269847.

14. Micek K, Hester KA, Chanda C, Darwar R, Dounebaine B, Ellis AS, et al. Critical success factors for routine immunization performance: A case study of Zambia 2000 to 2018. Vaccine: X. 2022:100166.

15. WGH. Subsidizing Global Health: Women’s unpaid work in Health Systems. 2022.

16. Pallas SW, Minhas D, Pérez-Escamilla R, Taylor L, Curry L, Bradley EH. Community health workers in low- and middle-income countries: what do we know about scaling up and sustainability? Am J Public Health. 2013;103(7):e74–82.

17. Bednarczyk R, Hester K, Dixit S, Ellis A, Escoffery C, Kilembe W, et al. Protocol: Identification and evaluation of critical factors in achieving high and sustained childhood immunization coverage in selected low- and lower-middle income countries 2021.

18. Exemplars. Exemplars in Global Health: Making Better Decisions in Global Health: Understand Positive Outliers to Inform Policy and Practice. 2021 [Available from: https://www.exemplars.health/.

19. Gottert A, McClair TL, Hossain S, Dakouo SP, Abuya T, Kirk K, et al. Development and validation of a multi-dimensional scale to assess community health worker motivation. J Glob Health. 2021;11:07008.

20. Agarwal S, Sripad P, Johnson C, Kirk K, Bellows B, Ana J, et al. A conceptual framework for measuring community health workforce performance within primary health care systems. Human resources for health. 2019;17(1):1–20.

21. Franco LM, Bennett S, Kanfer R. Health sector reform and public sector health worker motivation: a conceptual framework. Soc Sci Med. 2002;54(8):1255–66.

22. Colvin CJ, Hodgins S, Perry HB. Community health workers at the dawn of a new era: 8. Incentives and remuneration. Health Research Policy and Systems. 2021;19(3):106.

23. O’Mara-Eves A, Brunton G, Oliver S, Kavanagh J, Jamal F, Thomas J. The effectiveness of community engagement in public health interventions for disadvantaged groups: a meta-analysis. BMC Public Health. 2015;15(1):129.

24. Rohman A, Eliyana A, Purwana D, Hamidah H. Individual and organizational factors’ effect on knowledge sharing behavior. Entrepreneurship and Sustainability Issues. 2020;8(1):38.

25. Pfadenhauer LM, Gerhardus A, Mozygemba K, Lysdahl KB, Booth A, Hofmann B, et al. Making sense of complexity in context and implementation: the Context and Implementation of Complex Interventions (CICI) framework. Implementation science. 2017;12(1):1–17.

26. Damschroder LJ, Aron DC, Keith RE, Kirsh SR, Alexander JA, Lowery JC. Fostering implementation of health services research findings into practice: a consolidated framework for advancing implementation science. Implementation science. 2009;4(1):1–15.

27. Sakas Z, Hester KA, Ellis AS, Ogutu EA, Rodriguez K, Bednarczyk RA, et al. Critical success factors for high routine immunization performance: A multiple case study analysis of Nepal, Senegal, and Zambia. medRxiv. 2022:2022.11.08.22282076.

28. Katigbak C, Van Devanter N, Islam N, Trinh-Shevrin C. Partners in health: a conceptual framework for the role of community health workers in facilitating patients’ adoption of healthy behaviors. American journal of public health. 2015;105(5):872–80.

29. Sripad P, McClair TL, Casseus A, Hossain S, Abuya T, Gottert A. Measuring client trust in community health workers: A multi-country validation study. J Glob Health. 2021;11:07009.

30. Khatri RB, Mishra SR, Khanal V. Female Community Health Volunteers in Community-Based Health Programs of Nepal: Future Perspective. Frontiers in Public Health. 2017;5.

31. Kristen Devlin KFE, and Tanvi Pandit-Rajani. Community Health Systems Catalog Country Profile: Senegal. Arlington, VA: Advancing Partners & Communities. 2016.

32. Cothran D. Senegal’s community-based health System model: structure, strategies, and learning. 2019.

33. Ensor T, Green C, Quigley P, Badru AR, Kaluba D, Kureya T. Mobilizing communities to improve maternal health: results of an intervention in rural Zambia. Bulletin of the World Health Organization. 2013;92:51–9.

34. Jacobs C, Michelo C, Moshabela M. Implementation of a community-based intervention in the most rural and remote districts of Zambia: a process evaluation of safe motherhood action groups. Implementation science : IS. 2018;13(1):74-.

35. Zulu C, Michelo C, Mubita-Ngoma C. Evaluation of Training and Implementation Program for Community-Based Child Growth Monitors and Promoters in Zambia. 2017;1:34–9.

36. Perry H. A brief history of community health worker programs. Developing and strengthening community health worker programs at scale: a reference guide and case studies for program managers and policymakers, USAID, MCHIP. 2013.

37. Phiri SC, Prust ML, Chibawe CP, Misapa R, van den Broek JW, Wilmink N. An exploration of facilitators and challenges in the scale-up of a national, public sector community health worker cadre in Zambia: a qualitative study. Human Resources for Health. 2017;15(1):40.

38. Council OE. Towards an active, fair and socially cohesive Europe. Report of high level task force on social cohesion. TFSC (2007); 2008.

39. Olaniran A, Madaj B, Bar-Zeev S, Banke-Thomas A, van den Broek N. Factors influencing motivation and job satisfaction of community health workers in Africa and Asia-A multi-country study. Int J Health Plann Manage. 2022;37(1):112–32.

40. Sialubanje C, Massar K, Horstkotte L, Hamer DH, Ruiter RAC. Increasing utilisation of skilled facility-based maternal healthcare services in rural Zambia: the role of safe motherhood action groups. Reproductive Health. 2017;14(1):81.

41. Kasteng F, Settumba S, Källander K, Vassall A. Valuing the work of unpaid community health workers and exploring the incentives to volunteering in rural Africa. Health Policy Plan. 2016;31(2):205–16.

42. Panjabi R. Community Health Workers Are Vital; Governments Should Be Paying Them. TIME. 2019.

43. Coffey C, Espinoza Revollo P, Harvey R, Lawson M, Parvez Butt A, Piaget K, et al. Time to Care: Unpaid and underpaid care work and the global inequality crisis: Oxfam; 2020.

44. Maes K. The Lives of Community Helth Workers: Local Labor and Global Health in Urban Ethiopia. New York Routledge 2016. 188 p.

45. Raven J, Wurie H, Idriss A, Bah AJ, Baba A, Nallo G, et al. How should community health workers in fragile contexts be supported: qualitative evidence from Sierra Leone, Liberia and Democratic Republic of Congo. Human Resources for Health. 2020;18(1):58.

46. Central C. Gender and Community Health Workers: Three focus areas for programme managers and policy makers 2018 [Available from: https://chwcentral.org/twg_article/gender-and-community-health-workers-three-focus-areas-for-programme-managers-and-policy-makers/.

47. Ballard M, Westgate C, Alban R, Choudhury N, Adamjee R, Schwarz R, et al. Compensation models for community health workers: Comparison of legal frameworks across five countries. Journal of global health. 2021;11:04010-.

48. Kane S, Kok M, Ormel H, Otiso L, Sidat M, Namakhoma I, et al. Limits and opportunities to community health worker empowerment: A multi-country comparative study. Soc Sci Med. 2016;164:27–34.

49. Lee H-Y, Oh J, Heo J, Abraha A, Perkins JM, Lee J-K, et al. Association between maternal literacy and child vaccination in Ethiopia and southeastern India and the moderating role of health workers: a multilevel regression analysis of the Young Lives study. Global Health Action. 2019;12(1):1581467.

50. Dixit SM, Sarr M, Gueye DM, Muther K, Yarnko TR, Bednarczyk RA, et al. Addressing disruptions in childhood routine immunisation services during the COVID-19 pandemic: perspectives from Nepal, Senegal and Liberia. BMJ Global Health. 2021;6(7):e005031.

51. Schleiff MJ, Aitken I, Alam MA, Damtew ZA, Perry HB. Community health workers at the dawn of a new era: 6. Recruitment, training, and continuing education. Health Research Policy and Systems. 2021;19(3):1–28.

